# Non-invasive thoracoabdominal mapping of post-oesophagectomy conduit function

**DOI:** 10.1101/2023.01.10.23284370

**Authors:** Tim Hsu-Han Wang, Ashraf Tokhi, Armen Gharibans, Nicholas Evennett, Grant Beban, Gabriel Schamberg, Chris Varghese, Stefan Calder, Cuong Duong, Greg O’Grady

## Abstract

**Introduction:** Oesophagectomy is a complex procedure performed for malignant and benign conditions. Post-oesophagectomy conduit dysfunction is common, which can occur for several reasons including conduit dysmotility. However, reliable tools for evaluating conduit motility are lacking. A non-invasive device for gastric electrical mapping was recently developed to evaluate gastric electrical activity and function. This study aimed to assess the feasibility of BSGM in the post-oesophagectomy stomach.

**Methods:** Oesophagectomy patients from Auckland, New Zealand, were recruited. The Gastric Alimetry System® (New Zealand) was employed, comprising a stretchable array (8×8 electrodes), a wearable Reader, and validated iOS app for symptom logging. The protocol comprised a 30-minute baseline, a meal challenge, then 4 hours of post-prandial recordings. Analysis encompassed Principal Gastric Frequency, BMI-adjusted amplitude, Gastric Alimetry Rhythm Index (indicating rhythm stability), meal response, and symptoms. Adverse events were recorded.

**Results:** 6 patients were recruited and gastric activity was successfully captured in all except one with the colonic interposition (negative control). Four patients showed abnormalities indicating post-operative gastric hypofunction: four with low or abnormal frequency (<2.65 cycles/min), three with low amplitude (<22μV), two with low GA-RI (<0.25) and one with a reduced meal response. One patient had significant symptoms (nausea, early satiation) who demonstrated marked hypomotility in all four of these domains. No adverse events occurred.

**Conclusion:** Gastric Alimetry is a safe and feasible technique to non-invasively assess gastric conduit motility following oesophagectomy. Parameters may need adjustment for post-surgical anatomy. Clinical studies assessing the role in diagnosis and therapy can be advanced.

## Introduction

Oesophagectomy is a complex procedure performed for malignant and benign conditions. Procedural variations exist, dependent on patient and disease factors, with the stomach typically being used for reconstruction. Post-oesophagectomy conduit dysfunction is common, including delayed gastric conduit emptying (DGCE) (∼30%), gastro-oesophageal reflux (∼80%), and other chronic symptoms without a mechanical cause (1, 2). Emerging evidence implicates abnormal gastric electrophysiology as a contributing factor (3-5).

Although conduit dysfunction is multifactorial, dysmotility is a common contributing mechanism. However, it is clinically challenging to distinguish patients with dysmotility as opposed to alternative causes for symptoms (e.g. obstruction or pyloric dysfunction), as current tests such as endoscopy, fluoroscopy, radionuclear imaging, and manometry have limited accuracy, and/or are invasive or involve radiation. A safe and accurate test is needed to reliably assess conduit motility to inform correct therapy.

Gastric Alimetry® (Alimetry, New Zealand) is a new non-invasive test to evaluate gastric electrophysiology and function at high resolution (HR), recently receiving regulatory approvals for clinical use (6). This technique has been extensively validated (3, 7, 8), and is being applied in medical disorders, but has yet to be used for post-operative patients. Thisstudy therefore evaluated the feasibility of applying Gastric Alimetry after oesophagectomy to assess conduit motility.

## Methods

Patients who underwent oesophagectomy in Auckland, New Zealand, within the past three years were invited to participate following ethical approval (AH1125). Patients were excluded if they were undergoing chemotherapy/radiotherapy, had no post-operative computed tomography (CT) scan, or suffered mechanical obstruction. Clinical data including operation notes, imaging, endoscopy, and histopathology were evaluated.

Gastric Alimetry was performed under a protocol adapted for oesophagectomy. This device comprises an HR stretchable electrode array (8×8 electrodes; 20 mm spacing; 196 cm^2^), a wearable Reader, validated iOS app for symptom logging, and a cloud-based reporting platform (**Figure 1**) (9-11). Patients were fasted for >6 hours before array placement as guided by gastric position on CT (**Figure 1A-E**). After a 30 minute baseline recording, patients consumed a 218kCal meal (100mL nutrient drink and half an oatmeal energy bar), followed by a 4-hour postprandial recording with concurrent symptom logging.

**Figure 1.**
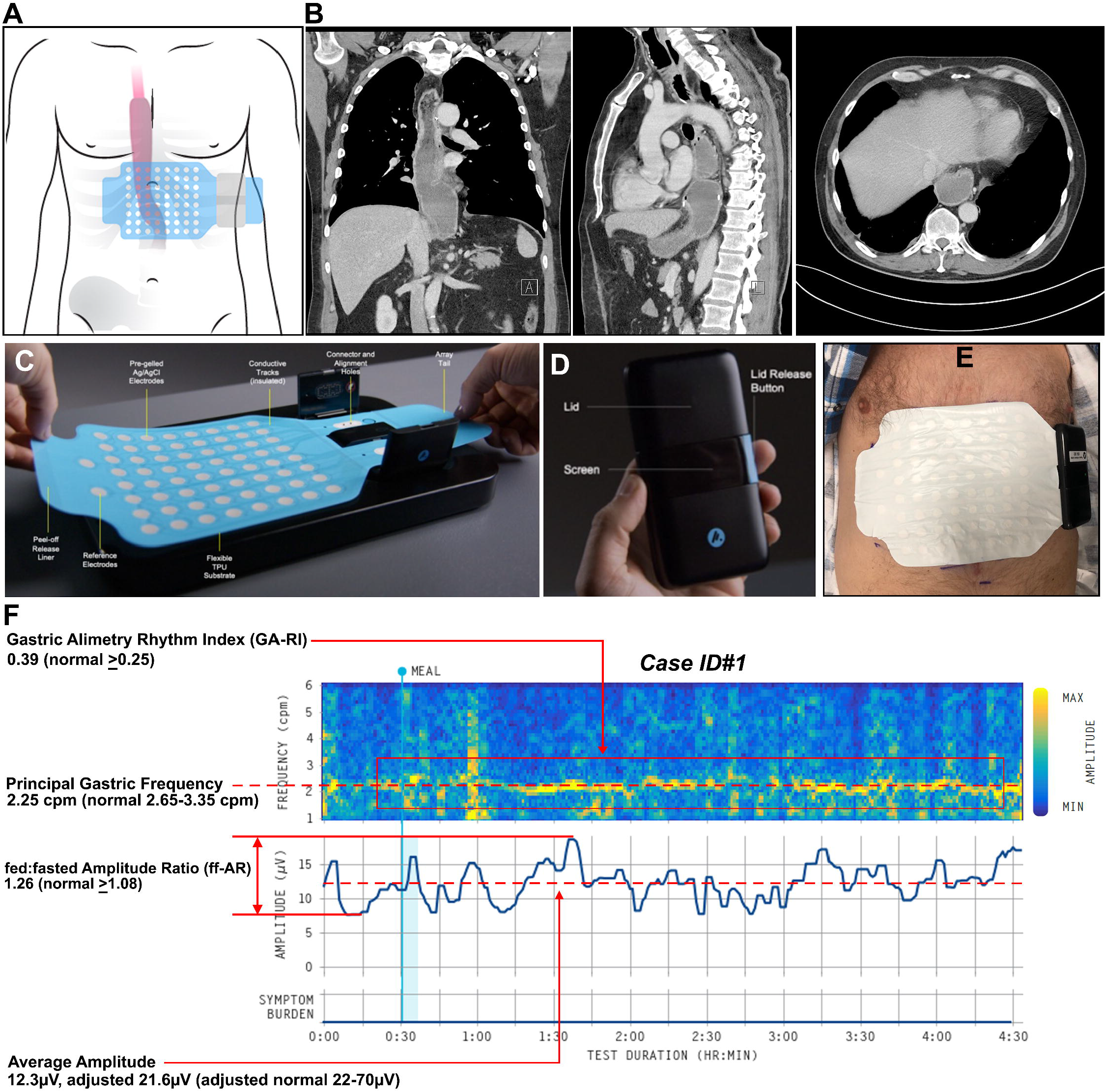
**A**. Array placement in relation to the thoraco-abdominal gastric conduit. **B**. Coronal, sagittal and axial views from CT scans of ID#1 illustrating the location of the stomach located in the posterior mediastinum, behind the heart, lungs, and liver. **C**. Gastric Alimetry stretchable electrode array. **D**. Gastric Alimetry wearable Reader. **E**. Gastric Alimetry electrode array and wearable Reader on the patient’s thoraco-abdominal region, per the placement depicted in **A. F**. Spectral map for Case ID#1 illustrating a reduced Principal Gastric Frequency and BMI-Adjusted Average Amplitude. Patient photo and imaging are used with patient’s written consent.

Spectral analysis was performed, encompassing four established metrics (12): Principal Gastric Frequency, BMI-Adjusted Amplitude, Gastric Alimetry Rhythm Index (GA-RI; reflecting pacemaker stability), fed:fasted Amplitude Ratio (ff-AR; indicating meal response with contractions), with comparison to reference intervals (13). Frequency was not reported if there was no rhythm (as measured by GA-RI) (10). Adverse events were recorded. Data were evaluated with descriptive statistics.

## Results

Demographic and operative data are reported in **Supplementary Table 1**. Six patients were recruited (all males; median age 65.5 years; range 58-73). Oesophagectomies were performed between 6.5 months - 3 years prior, with the standard procedure including vagotomy and pyloroplasty. Indications were cancer (n=4), Barrett’s oesophagus (n=1) and achalasia (n=1). One case (ID#6) developed a necrotic gastric conduit prompting resection, formation of cervical oesophagostomy and feeding jejunostomy on day 6 following surgery, with subsequent colonic interposition graft with Roux-en-Y reconstruction 8 months later. This case served as a negative control.

All patients except one were largely asymptomatic at time of testing. The symptomatic patient (ID#5) reported moderate to severe nausea, vomiting, early satiation, abdominal pain, reflux, and a poor quality of life.

Gastric activity was successfully captured non-invasively in all cases (**Figures 1, 2**). Four cases (IDs#1-2,4-5) had at least one abnormal parameter, all showing reduced motility profiles (**Figure 1F, Figure 2A,C-E**). Of these, low or abnormal frequency was the most common abnormality (4/4 cases), followed by low amplitude in 3/4, low GA-RI in 2/4 and low ff-AR in 1/4 (**Figure 2E**). The symptomatic patient (ID#5) was found to have abnormalities in all four domains, with symptoms being maximal when activity was weakest (**Figure 2D,E)**. ID#3 was the only case with normal parameters throughout (**Figure 2B,E**), who had minimal gastric resection (<4 cm).

**Figure 2.**
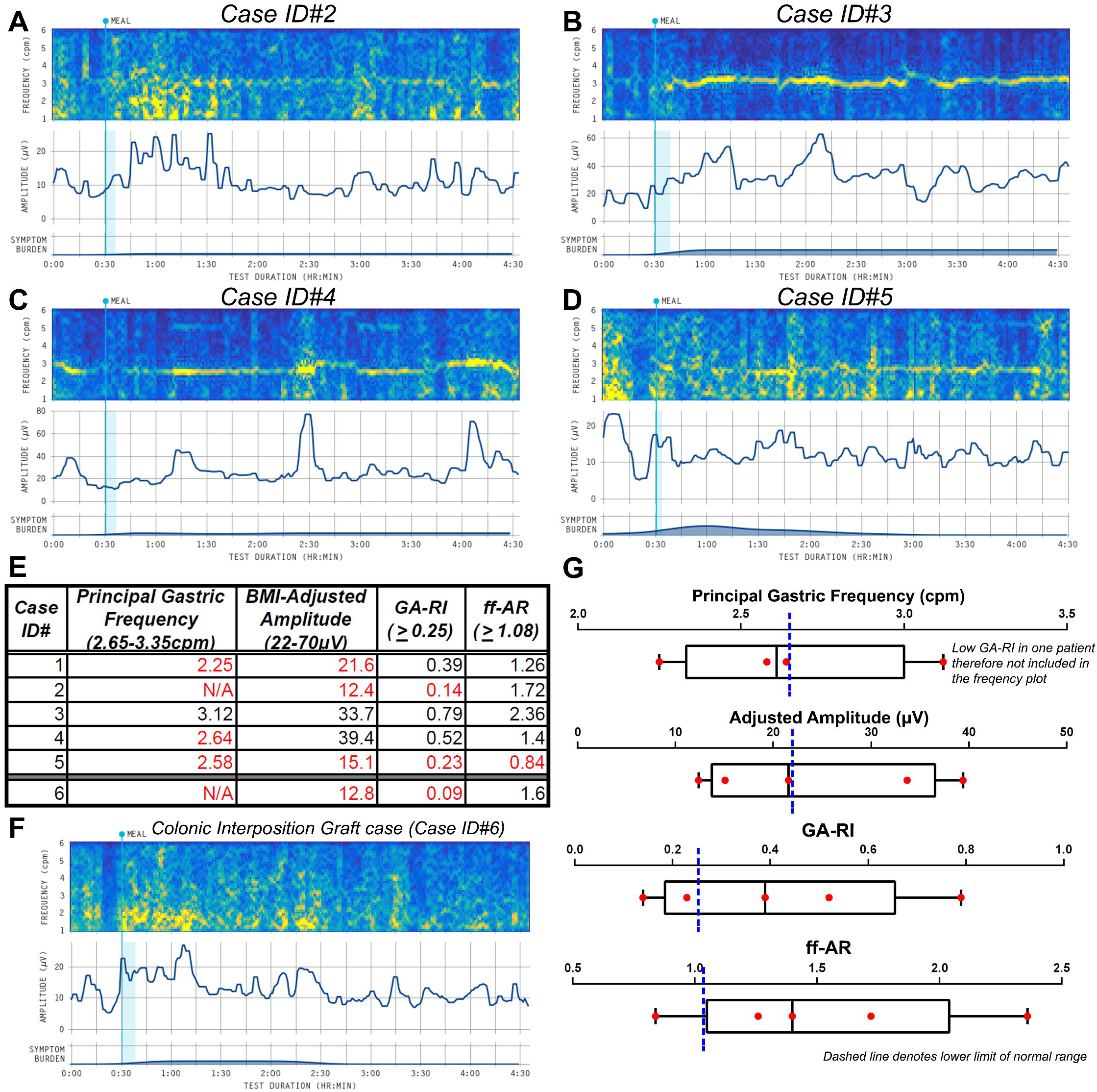
**A-D** illustrates case ID#2 to ID#5’s spectral maps with associated symptom burden plots. Quantitative analysis is presented in **E** with reference intervals as shown. Quantitative results for case ID#1 shown in **Figure 1F** is also presented in **E. F**. Spectrogram for the patient with the colonic interposition graft. **G**. Box and whiskers graph for the quantitative results for case ID#1 to ID#5. The blue dashed line represents the lower limit of the reference interval for each Gastric Alimetry spectral metric.

In the negative control (ID#6), no gastric activity was identified, but low frequency burst activity was evidence consistent with colonic activity immediately post-prandially (**Figure 2F**) (14).

No adverse reactions occurred.

## Discussion

Persistent upper gastrointestinal symptoms in the absence of mechanical obstruction are common after oesophagectomy. Contributing factors include conduit dysmotility, hypersensitivity/pain syndromes, dumping syndrome, and pyloric dysfunction, which may overlap and are difficult to differentiate on clinical history and current tests. This study shows the safety and feasibility of a new test called Gastric Alimetry for non-invasively evaluating the function of the deep-seated post-oesophagectomy gastric conduit.

Gastric surgery modifies the electrical conduction system that coordinates contractions (15), with previous studies implicating abnormal electrophysiology in conduit dysfunction (3-5). However, reliable techniques to assess conduit function have been lacking. Recent advances have enabled substantial progress in evaluating gastric electrophysiology in health and disease (7, 16, 17). A legacy technique termed electrogastrography (EGG) previously attempted to capture gastric electrical activity from the skin surface, but was limited by low resolution and high sensitivity to noise (5). Gastric Alimetry overcomes these problems by employing an HR array together with sophisticated signal processing algorithms (9, 10), that were shown effective even with conduits positioned in the thorax and posterior mediastinum. The patient with a total gastrectomy and colonic interposition graft served as a negative control, further increasing confidence in the current findings.

Previously, Gastric Alimetry has been exclusively performed in patients with normal gastric anatomies, in whom reference ranges were developed (12, 13, 17). Some adjustments to interpretations will therefore be required as the test is applied to post-operative patients. Specifically, normative values for amplitude will need to be re-defined due to the greater distance between the stomach and the array, and this work is currently in progress. Additionally, meal sizes were reduced by 50% vs the standard Gastric Alimetry test to account for the reduced gastric remnant volume, which is considered adequate to stimulate gastric activity (6).

Reduced motility parameters were the dominant finding in this post-oesophagectomy cohort, observed as reductions in frequency, rhythm stability (GA-RI) and meal responses (ff-AR), except for one patient who had a minimal gastric resection. Reduced frequency likely reflects resection of the native gastric pacemaker, leading to the development a new lower-frequency pacemaker (18). Low GA-RI likely reflects gastric neuromuscular dysfunction due to aberrant pacemaker recovery (6), while reduced meal responses could reflect loss of vagal input (15). While vagotomy is inevitable to allow lymph node harvest in cancer patients, evolving techniques offer vagal-sparing esophagectomies for non-malignant indications (eg. achalasia, or stricturing disease) may assist in avoiding the vagotomy-associated complications (19).

Validated symptom profiling is also performed with the Gastric Alimetry test. While detailed symptom analysis was not a focus of this feasibility study, symptom profiling is proving useful elsewhere in distinguishing cases with hypersensitivity and pain syndromes, particularly when gastric function is normal, and is likely to be valuable in future post-operative studies (6, 11). Emerging spatial mapping techniques will also allow determination of electrical propagation patterns in future studies (9).

With feasibility established, future studies can now be conducted applying this technique on larger cohorts of patients after oesophagectomy. Such work will enable improved characterisation of pathophysiology and symptom correlations, in order to guide therapeutic decisions, as is being performed in other gastric disorders (6). In addition, the new test is also now being evaluated for its potential in gastric dysfunction after pancreaticoduodenectomy (20).

## Data Availability

All data produced in the present work are contained in the manuscript

## Acknowledgements

We thank the volunteers who participated in this research, the Auckland City Hospital clinical nurse specialist Elaine Yi and our Auckland clinical research coordinators Gen Johnston and India Wallace. Research preregistration was not completed.

