## Supplementary Table 1 for "Non-invasive thoracoabdominal mapping of post-oesophagectomy conduit function"

| ID# | Age group  (years) | Gender | Procedure | Indication | Amount of stomach resected | BMI | Interval between surgery and mapping (months) |
| --- | --- | --- | --- | --- | --- | --- | --- |
| 1 | 55-60 | M | Hybrid Ivor-Lewis oesophagectomy and left adrenalectomy | Achalasia | 9.5cm x 4.5cm x 3.5cm of stomach resected | 28 | 14 |
| 2 | 60-65 | M | Minimally invasive three-stage oesophagectomy | Oesophageal cancer | 14cm greater curvature,  5.5cm lesser curvature | 22.3 | 17 |
| 3 | 65-70 | M | Hybrid Ivor-Lewis oesophagectomy | Oesophageal cancer | 4cm greater curvature,  4cm lesser curvature | 22.9 | 36 |
| 4 | 60-65 | M | Minimally invasive oesophagectomy | Barrett's oesophagus with high grade dysplasia | 4.5cm lesser curvature | 30.4 | 6.5 |
| 5 | 65-70 | M | Minimally invasive three-stage oesophagectomy | Oesophageal cancer | 13cm greater curvature | 23.2 | 15.5 |
| 6 | 70-75 | M | 1. Hybrid two-stage oesophagectomy 2. Colonic interposition graft | Oesophageal cancer | 20cm greater curvature,  12.5cm lesser curvature | 21.7 | 12 |
